# Extremely Preterm Children Demonstrate Interhemispheric Hyperconnectivity During Verb Generation: a Multimodal Approach

**DOI:** 10.1101/2020.10.30.20222448

**Authors:** Maria E. Barnes-Davis, Stephanie L. Merhar, Scott K. Holland, Nehal A. Parikh, Darren S. Kadis

## Abstract

Children born extremely preterm (EPT, <28 weeks gestation) are at risk for delays in development, including language. We use fMRI-constrained magnetoencephalography (MEG) during a verb generation task to assess the extent and functional connectivity (phase locking value, or PLV) of language networks in a large cohort of EPT children and their term comparisons (TC). 73 participants, aged 4 to 6 years, were enrolled (42 TC, 31 EPT). There were no significant group differences in age, sex, race, ethnicity, parental education, or family income. There were significant group differences in expressive language scores (p<0.05). Language representation was not significantly different between groups on fMRI, with task-specific activation involving bilateral temporal and left inferior frontal cortex. There were group differences in functional connectivity seen in MEG. To identify a possible subnetwork contributing to focal spectral differences in connectivity, we ran Network Based Statistics analyses. For both beta (20-25 Hz) and gamma (61-70 Hz) bands, we observed a subnetwork showing hyperconnectivity in the EPT group (p<0.05). Network strength was computed for the beta and gamma subnetworks and assessed for correlation with language performance. For the EPT group, exclusively, strength of the subnetwork identified in the gamma frequency band was positively correlated with expressive language scores (r=0.318, p<0.05). Thus, interhemispheric hyperconnectivity is positively related to language for EPT children and might represent a marker for resiliency in this population.

## 1. INTRODUCTION

The development of noninvasive neuroimaging has enabled assessment of language representation across the lifespan, with task-based functional magnetic resonance imaging (fMRI, the gold standard for language mapping) demonstrating bilateral patterns of language activation which become increasingly left lateralized for most children as they mature into adulthood (Holland et al., 2001; Holland et al., 2007; Wood et al., 2004). Language relies on fast neuronal processes which can be transient and brief, occurring in a matter of milliseconds, and—by characterizing fast oscillatory activity--we can map language (Benasich, Gou, Choudhury, & Harris, 2008; Doesburg, Tingling, MacDonald, & Pang, 2016; Kadis, Dimitrijevic, Toro-Serey, Smith, & Holland, 2016; Kadis et al., 2007; Kadis et al., 2011). Magnetoencephalography (MEG) though less widely used than fMRI, affords sub-millisecond temporal resolution and can localize sources in the cortex, especially when combined with MRI (Kadis et al., 2016). This makes MEG especially suited to lifespan assessment of developing language networks in typically developing children and in clinical populations at risk for atypical language development (Barnes-Davis, Merhar, Holland, & Kadis, 2018; Gaudet, Hüsser, Vannasing, & Gallagher, 2020; Taylor, Donner, & Pang, 2012).

Prematurely born children represent one such clinical population. Preterm birth is a public health crisis, impacting 10% of children born in the United States every year and up to 15 million children globally every year (Blencowe et al., 2013; Martin, Hamilton, Osterman, & Driscoll, 2019). With advances in perinatal and neonatal care, children born as preterm as 22 weeks are now surviving. However, they still have significant risk for sensory and neurodevelopmental impairment (Adams-Chapman et al., 2018; Barre, Morgan, Doyle, & Anderson, 2011; Hutchinson, De Luca, Doyle, Roberts, & Anderson, 2013; Mikkola et al., 2005; Moore, Lemyre, Barrowman, & Daboval, 2013; van Noort-van der Spek, Franken, & Weisglas-Kuperus, 2012; Vohr, 2014). Children born preterm are at an increased risk for language impairment versus their term peers, with up to 40% of preterm children experiencing language delays and approximately 20% receiving a diagnosis of specific language impairment (Barre et al., 2011; Foster-Cohen, Edgin, Champion, & Woodward, 2007; Foster-Cohen, Friesen, Champion, & Woodward, 2010; van Noort-van der Spek et al., 2012; Woods, Rieger, Wocadlo, & Gordon, 2014). Term equivalent age neuroimaging with cranial ultrasound or structural MRI and early language testing such as the Bayley Scales of Infant Development at 18-24 months corrected gestational age leave a large proportion of the variance in later language performance unaccounted for and do not accurately predict delays (Luttikhuizen dos Santos, de Kieviet, Konigs, van Elburg, & Oosterlaan, 2013; Woods et al., 2014). This is important because early intervention programs are one of the few therapies that can improve neurodevelopmental outcomes, including language performance. Addressing language impairment early on might mitigate later impact on scholastic attainment and quality of life (Barre et al., 2011; Vieira & Linhares, 2011; Vohr, 2014).

Despite the numerous prenatal and neonatal risk factors for impairment that preterm children experience, there is great variability in outcome, with some children born extremely preterm (less than 28 weeks completed gestation) doing well (Garfield, Karbownik, Murthy, & et al., 2017). Furthermore, adults who were born preterm and their families express frustration that they were given overwhelming lists of risk factors in the neonatal intensive care unit (NICU) but little to no information on how to counter these risks (Leavy, Prina, Martinez Caceres, & Bauer, 2015; Saigal, 2014). Investigations of resiliency factors—those that confer a positive outcome despite risk—should be prioritized along with risk factors and factors that are universally protective. The search for functional neuroimaging markers of risk and resiliency in the context of preterm birth is important in that it could provide metrics for risk stratification and predictive markers for later language functioning, perhaps before children exhibit verbal or language skills. Additionally, through the investigation of aberrant developmental trajectories (including those of children who spend the third trimester of gestation ex utero) we might gain new insights regarding the neurobiology of language that could be of utility for other clinical groups and for typically developing children. Existing literature evaluating connectivity supporting language in preterm children to date has relied primarily on resting state and task-based functional MRI. Taken as a whole, these functional MRI studies report increased interhemispheric connectivity and increased involvement of right-sided language homologue areas (Barnes-Davis et al., 2018; Choi, Vandewouw, Young, & Taylor, 2018; Gozzo et al., 2009; Kwon et al., 2015; Kwon et al., 2016; Myers et al., 2010; Schafer et al., 2009; Scheinost et al., 2015; Wilke, Hauser, Krageloh-Mann, & Lidzba, 2014). However, relationships between connectivity and inconsistent.

MEG has only been used in a handful of studies investigating neural networks in children born preterm (Barnes-Davis et al., 2018; Doesburg, Moiseev, Herdman, Ribary, & Grunau, 2013; Doesburg, Ribary, Herdman, Miller, et al., 2011; Doesburg, Ribary, Herdman, Moiseev, et al., 2011; Mossad, Smith, Pang, & Taylor, 2017; Sato et al., 2019; Ye, AuCoin-Power, Taylor, & Doesburg, 2016). Our prior pilot study of 15 children born EPT and 15 term comparison children (TC) was the first to use MEG to investigate the connectivity of language networks in prematurity. We assessed representation and connectivity of language networks at 4-6 years of age using multimodal imaging during a receptive language paradigm (passive stories listening task). Language representation (fMRI) was not significantly different between groups (Barnes-Davis et al., 2018). However, functional connectivity (MEG) was significantly increased in EPT, with significantly greater network strength than term controls TC. Connectivity was increased in bilateral temporal regions, in accordance with fMRI connectivity studies in older preterm participants (Gozzo et al., 2009; Scheinost et al., 2015). Prior studies could not conclude if hyperconnectivity in EPT reflects an alternate developmental trajectory or immaturity of normal language lateralization (Gozzo et al., 2009). We were able to leverage the temporal resolution of MEG to assess information flux and concluded that hyperconnectivity was due to increased drivers in right perisylvian cortex, supporting our theory of an alternate developmental trajectory and not merely a delay in normal language lateralization (Barnes-Davis et al., 2018). Additionally, through investigation of diffusion imaging obtained in the same study visit, we were able to show this increased connectivity was due to an atypically robust extracallosal pathway connecting bilateral perisylvian cortices (Barnes-Davis, Williamson, Merhar, Holland, & Kadis, 2020b). Performance on language tasks was positively correlated with extracallosal structural connectivity for the preterm group exclusively.

Prior MEG studies suggest that activation in different frequency bands or involvement of different language regions in typically developing children could be due to differences in language processing based on participant characteristics or differences in task characteristics (expressive versus receptive, etc.) (Gaudet et al., 2020). Thus, it is important to consider clinical groups, task demands, and language modalities when interpreting electrophysiological studies of language networks in the brain. MEG studies including preterm children have reported decreased task-based and resting state functional connectivity on the whole brain level (Doesburg et al., 2013; Doesburg, Ribary, Herdman, Miller, et al., 2011; Doesburg, Ribary, Herdman, Moiseev, et al., 2011; Ye et al., 2016). However, none have focused on language and some did not significantly correlate connectivity metrics with performance. The aim of this current paper is to investigate functional connectivity during an expressive language paradigm (a widely used verb generation task) to assess the extent and connectivity of language networks in the largest cohort of children born extremely preterm followed by MEG (known to us) to date. Furthermore, we will compare connectivity of EPT children to that of their term counterparts and assess relationships with performance. We hypothesize that EPT children will have increased bitemporal functional connectivity versus TC and that this hyperconnectivity will be positively related to performance on standardized language assessments (particularly expressive language tasks) for EPT but not for TC.

## 2. METHODS

### 2.1. Participants

This is a cohort study with 73 participants aged 4 to 6 years recruited from the greater Cincinnati area. TC (n=42) were recruited from local pediatric clinics and community announcements. EPT children (n=31) were recruited from Cincinnati-area neonatal intensive care units (NICUs) if they were born at less than 28 weeks gestation and were without parenchymal lesions, hemorrhage, or IVH >grade 2 on neonatal cranial ultrasound. Children with cerebral palsy, seizures, migraines, and formally diagnosed psychiatric disorders such as autism or ADHD were excluded from both groups. Children with contraindications for MRI or MEG scanning (metal implants, insulin pumps, etc.) were excluded from both groups. Imaging and behavioral data were included from 3 studies investigating the language development of term born and preterm children using multimodal neuroimaging. All 3 studies are approved by the Cincinnati Children’s Hospital Medical Center IRB and the IRBs of Cincinnati-area delivery hospitals. All study procedures conformed to the US Federal Policy for the Protection of Human Subjects. Written informed consent was obtained from parents and verbal assent from all children.

### 2.2. Demographic and Neuropsychological Assessments

For all participants, demographic data were recorded, including age, sex, race, and ethnicity. Socioeconomic variables such as parental education and family income were also recorded by the parents. Participants completed a neuropsychological testing battery including the Peabody Picture Vocabulary Test 4 (PPVT); the Expressive Vocabulary Test 2 (EVT); and the Weschler Nonverbal Test of Abilities (WNV). The PPVT (Dunn, Dunn, & Lenhard, 2015) is a well-normed and well-validated test of receptive language/vocabulary that has been widely used in studies of outcomes in prematurity (Luu et al., 2009; Ment et al., 2006; Mullen et al., 2011; Myers et al., 2010). The EVT (Williams, 2007) is co-normed with the PPVT and is a well validated measure of expressive language. The WNV (Wechsler & Naglieri, 2006) is a nonverbal test of general abilities especially developed for populations where there is a high risk of language impairment or where English might not be the primary spoken language.

### 2.3. Verb Generation Task

The same verb generation stimuli were used in both MEG and fMRI. For fMRI, they were presented in a block design; for MEG, they were presented in an event-related, random presentation. This auditory covert verb generation task has been reported previously by our group (Holland et al., 2001; Kadis et al., 2016; Youssofzadeh, Williamson, & Kadis, 2017). In brief, the task consisted of nouns spoken by a female voice (verb generation condition) or speech-shaped noise, matched to the noun stimuli for spectral content and amplitude envelope (noise condition). Children were asked to covertly generate a verb or “think of an action word” upon hearing nouns; in contrast, they were asked to quietly attend to the noise stimuli, without responding. Prior to MEG recording, participants were trained on an overt version of the task to assess understanding and promote compliance during acquisition. During MEG acquisition, 71 distinct nouns and 71 noise trials were randomly presented. During fMRI acquisition, the noun stimuli were presented at a rate of once every 5 sec, in five 30-sec blocks. During the control block, children were asked to simply listen to the noise stimuli, presented every 5 seconds, in six 30-sec blocks. Participants listened to a total of 30 nouns and 36 noise tones during the fMRI recordings, lasting less than 6 minutes.

### 2.4. Magnetoencephalography Acquisition

To avoid potential magnetization from the MRI, MEG was always acquired before MRI imaging on a 275-channel whole-head CTF system (MEG International Services Ltd., Coquitlam, BC). Neuromagnetic activity was recorded at 1200Hz. Subjects were tested while awake and in the supine position. Auditory stimuli were presented via a calibrated audio system comprised of distal transducer, tubing, and ear inserts (Etymotic Research, IL, USA). Head localization coils were placed at nasion and preauricular locations to monitor movement throughout the recording. Following acquisition, the coils were replaced with radio-opaque markers, to facilitate offline co-registration of MRI and MEG data.

### 2.5. Magnetic Resonance Acquisition

All subjects were studied awake without sedation in the presence of an experienced pediatric radiology technician, a pediatric clinical research coordinator, and a parent or legal guardian. This sample consists of children from 2 distinct studies on multimodal imaging of language in prematurity. The first study used a single echo fMRI acquisition. The second study used a multi-echo fMRI acquisition. For both studies, extremely preterm and term children were enrolled and the task, imaging site, and scanner remained the same. We conducted fMRI scanning at 3T (Philips Achieva scanner). The single echo acquisition (used for 12 EPT children and 13 TC children) had TR / TE = 2000 / 30 ms, flip=75°, 2.8 x 2.8 x 3.0 mm voxels. The multi echo acquisition (used for 17 EPT children and 29 TC) had TR 1381.14 ms; TE 14, 32, and 50 ms; 3.0 x 3.0 x 3.0 mm voxels; 308 dynamics; multiband factor 3; and in-plane SENSE factor of 3). For this analysis, we used only the middle echo (32 ms) for fMRI preprocessing and first and second level contrasts. Scan duration for both acquisitions was about 5 minutes. We obtained 3D T1-weighted volumes at 1.0 x 1.0 x 1.0 mm.

### 2.6. Demographic and Neuropsychological Data Analyses

Between group comparisons of continuous variables (age, assessment scores) were performed using independent samples t-tests in SPSS Statistics 26 running in Mac OS 10.15.6. Categorical variables (sex, race, ethnicity, parental education, household income) were compared between groups using Fisher’s exact test.

### 2.7. Magnetic Resonance Imaging Analyses

Task-based fMRI data were analyzed using a conventional general linear model preprocessing pipeline in SPM12 (http://www.fil.ion.ucl.ac.uk/spm/software/spm12/) running in MatLab R2020a (MathWorks Inc., Natick, MA). This has been detailed in previous publications (Barnes-Davis et al., 2018; Kadis et al., 2016). The preprocessing pipeline included realignment, co-registration to the individual subject’s T1-weighted images, normalization to template space (MNI 152), and smoothing with a full width half maximum (FWHM) of 5 mm. Task-based fMRI data were subjected to first- and second-level analyses as detailed in previous publications (Barnes-Davis et al., 2018; Kadis et al., 2016). At the first level, a Verb minus Noise contrast, with size motion parameters included as nuisance regressors, was evaluated to assess activation that was measured in response to the language stimulus versus the noise condition. At the second level, first level condition contrasts were collected and subjected to a between groups independent samples t test evaluating the spatial distribution of activation in response to the language task. For this contrast, p<0.001 and k=8. This was assessed with and without a family-wise error correction of p<0.05 and with and without accounting for imaging acquisition (single echo versus multi echo). Groupwise activation, including all preterm and term participants, were included in the joint activation map. This was parcellated into 200 random units (Craddock, James, Holtzheimer, Hu, & Mayberg, 2012). Parcels having greater than 10% active voxels were retained, and the center of gravity was calculated for each parcel. These coordinates were determined and served as coordinate for virtual sensors in MEG connectivity analyses and subsequent graph theoretical analyses.

### 2.8. Magnetoencephalography Analyses

#### 2.8.1. Virtual Sensor Extraction and Preprocessing

Data were analyzed using FieldTrip, an open-source MatLab toolbox for analyses of electrophysiological data, including MEG (Oostenveld, Fries, Maris, & Schoffelen, 2011). Continuous neuromagnetic data were bandpass filtered from 0.1Hz to 100Hz, and 60Hz power-line noise attenuated using a sharp discrete Fourier transform filter. The data were then segmented into epochs of interest (700-1200ms from onset of language stimuli) to remain consistent with previous work in term children (Kadis et al., 2011; Youssofzadeh et al., 2017). Scanner jump artifacts were automatically detected, and the affected trials were rejected. We required at least 90% trial retention for inclusion in subsequent analyses. Realistic single-shell head models were constructed from the participants’ 3D T1 weighted images (Nolte, 2003). A 3-D grid was constructed with dipole resolution of 5 mm on a template (MNI151) brain volume. The template grid was warped to individual anatomical space, and used to develop individual source models. Using a linearly constrained minimum-variance beamformer (LCMV) with 0.1% regularization, we estimated the time series of activity at each network node (virtual sensor analysis) similar to our previously reported work (Barnes-Davis et al., 2018).

#### 2.8.2. Functional Connectivity

Phase locking value (PLV) was used to assess functional connectivity in MEG. PLV reflects the consistency of a phase relationship between two signals, with values ranging from 0 (no phase synchrony) to 1 (perfect phase synchrony) (Bastos & Schoffelen, 2016; Lachaux, Rodriguez, Martinerie, & Varela, 1999; Youssofzadeh et al., 2017). Using FieldTrip, connectivity analyses were performed on the time courses at each virtual sensor. The trials were zero padded to 2 seconds total to minimize edge effects. A multi-taper method FFT (+/- 5Hz smoothing) decomposition, over a frequency range of 2-70 Hz and a step of 0.5 Hz, was used. The connectivity spectra were then plotted by group and visually inspected. Frequency bands in which there were apparent group differences were identified; connectivity within those bands was averaged, and the resulting adjacency matrices were subjected to analysis using Network Based Statistics (NBS). The approach is useful for identifying potential subnetworks contributing to observed network-wide group differences in connectivity. Here, we studied network extent as a function of group across a range of initial thresholds, using an overall p value of 0.05 and for 5000 permutations (Zalesky, Fornito, & Bullmore, 2010). For the purpose of visualization, the median t-value yielding significant NBS results was used to characterize the subnetworks showing group differences. Surviving connections were summed for each subject to assess network strength. Network strength was then assessed for correlation with expressive language scores across and within groups.

## 3. RESULTS

### 3.1. Demographic and Neuropsychological Assessments

There were no significant group differences in age, sex, race, ethnicity, highest parental education, combined family income, PPVT scores, or WNV scores (see Table 1). There were significant group differences in expressive language scores (mean standardized EVT scores were 102.6 +/- standard deviation of 9.4 for the EPT group and 111.7 +/- standard deviation of 15.9 for the TC group). An outlier was identified with significantly decreased scores (greater than 2 standard deviations below the mean) on standardized assessments (population mean for each of these instruments is 100, with a standard deviation of 15). This individual was removed from subsequent analyses, leaving 30 EPT children and 42 TC children in the final connectivity analysis. For the preterm group, gestational age at birth ranged from 24 weeks to 27 weeks and 5 days. For the term group, gestational age ranged from 37 to 41 weeks.

**Table 1:** Demographics and Neuropsychological Data for Entire Sample.

|  |  | **Preterm**<br>**(n=31)** | **Term**<br>**(n=42)** | **p Value** |
| --- | --- | --- | --- | --- |
| **Age (Years, Mean ± SD)** |  | 5.68 ± 0.84 | 5.62 ± 0.95 | 0.79 |
| **Gestational Age (Weeks + Days)** |  | 26+0 | 39+2 | <0.01 |
| **Sex** | Females | 17 | 22 | 1 |
|  | Males | 14 | 20 |  |
| **Race** | White/Caucasian | 19 | 29 | 0.79 |
|  | Black/African American | 9 | 8 |  |
|  | Other/Multiple | 1 | 1 |  |
|  | No Response | 2 | 4 |  |
| **Ethnicity** | Hispanic/Latino/Latina | 3 | 1 | 0.39 |
|  | Not Hispanic/Latino/Latina | 27 | 38 |  |
|  | No Response | 1 | 3 |  |
| **Family Income** | < \$25000 | 5 | 4 | 0.69 |
| | \$25000-\$50000 | 3 | 7 | |
| | \$50000-\$75000 | 10 | 8 | |
| | \$75000-\$100000 | 4 | 5 | |
| | \$100000-\$150000 | 4 | 10 | |
| | >\$150000 | 4 | 5 | |
|  | No Response | 1 | 3 |  |
| **Parental Education** | High School Diploma | 3 | 4 | 0.35 |
|  | Some College | 11 | 9 |  |
|  | College Diploma | 4 | 2 |  |
|  | Post Graduate | 12 | 24 |  |
|  | No Response | 1 | 3 |  |
| **Receptive Language** | PPVT-4 (Mean ± SD) | 113 ± 11.0 | 119 ± 18 | 0.08 |
| **Expressive Language** | EVT-2 (Mean ± SD) | 102 ± 9 | 112 ± 16 | <0.01 |
| **General Abilities** | WNV (Mean ± SD) | 103 ± 15 | 109 ± 16 | 0.13 |

Note: Categorical variables were tested using Fisher's Exact Test and p values are reported. Continuous variables were tested using t tests and p values are reported with 95% confidence intervals.
CI = Confidence Interval. SD = Standard Deviation. PPVT-4 = Peabody Picture Vocabulary Test. EVT-2 = Expressive Vocabulary Test. WNV = Wechsler Non-Verbal Scale of Ability.

### 3.2. Activation in Functional Magnetic Resonance Imaging

Of the 73 children included in the study, 7 TC children and 4 EPT children did not complete neuroimaging due to scheduling issues (e.g. arriving late to the study visit and needing to stop visit before all tasks were completed) or noncompliance with task (movement or wanting to stop imaging). Thus, 62 children were included in the fMRI analyses (35 TC and 27 EPT).

There were no significant differences in language representation in fMRI, between groups. Activation in response to verb generation versus the noise condition involved the expected regions, including bilateral temporal and left inferior frontal cortex (Figure 1A). To create a balanced joint activation map for subsequent MEG analyses, a subset (n = 53) of participants were randomly selected for participation in the mask, with equitable representation between groups (EPT versus TC) and study acquisitions (single echo versus multi-echo fMRI). The joint activation map was generated from the first level contrast maps (verbs versus noise) from 14 TC from the multi-echo acquisition, 14 EPT from the multi-echo acquisition, 13 TC from the single echo acquisition, and 12 EPT from the single echo acquisition.

**Figure 1:**
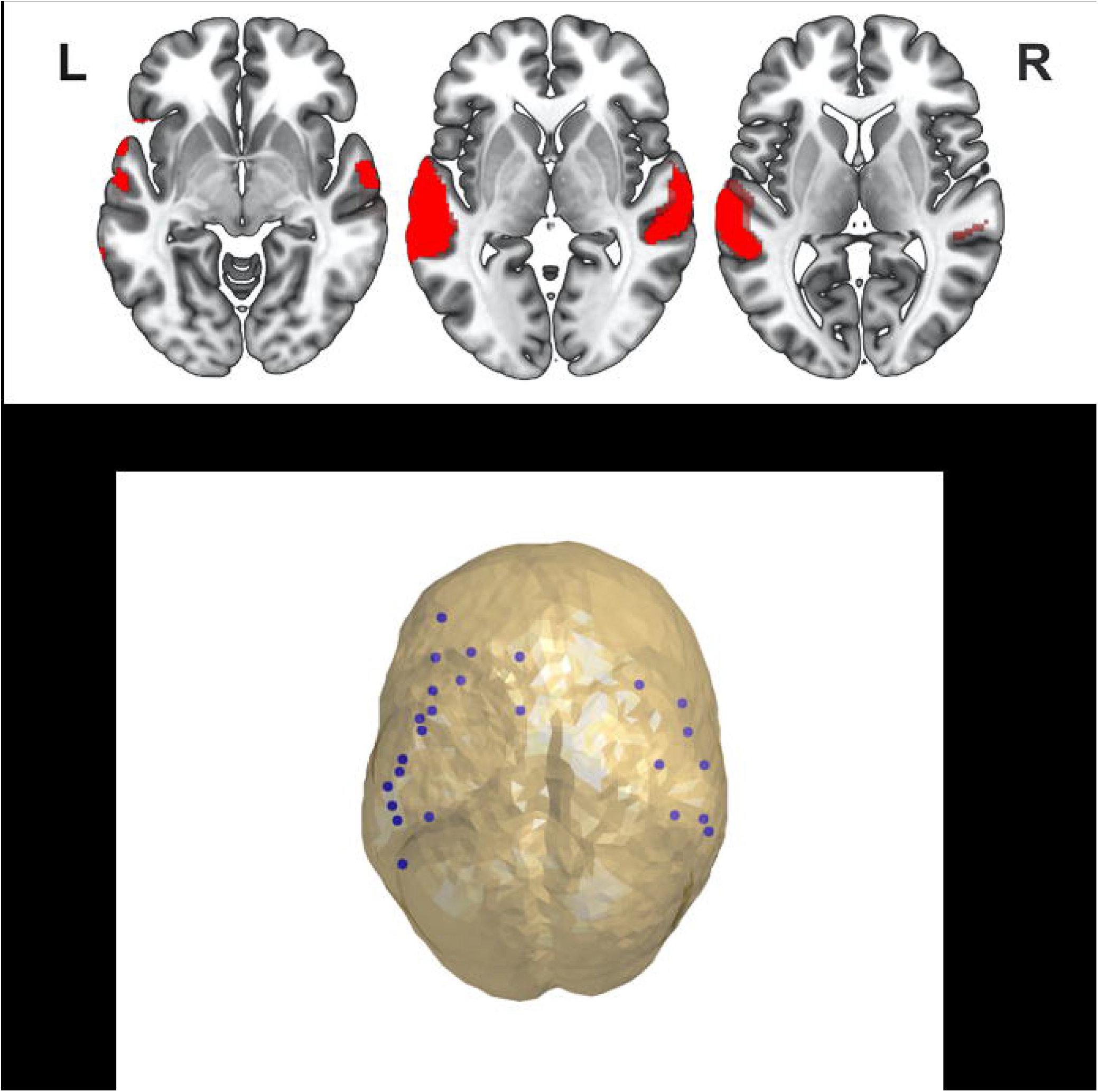
Functional MRI activation maps and resultant network nodes. A) Three slices through temporal, parietal, and left frontal language areas showing general linear model (GLM) analysis across all participants (EPT + TC) with typical bilateral activation in response to language stimuli (auditorily presented nouns for verb generation) versus noise condition (*p* < 0.001, k = 8) and 3D surface rendering of the normalized brain averaged across all participants to show bilateral activation from a sagittal perspective. B) The initial activation map across the entire sample is subjected to a 200-unit parcellation scheme to produce the “nodes” for network analysis, shown in blue.

### 3.3. Connectivity in Magnetoencephalography

Virtual sensors were obtained from multiplying the fMRI verb generation joint activation map with a 200 unit random parcellation map and extracting center of gravity coordinates from parcels with over 10% active voxels. These virtual sensors are shown in in Figure 1B and served as nodes for subsequent connectivity and network analyses. Phase locking value (PLV) was computed for all node pairs for each participant as described above, and plotted as a function of frequency, for each group (Figure 2). We visually inspected the connectivity spectra and identified 2 bands in which we suspected EPT and TC children would significantly differ. We noted apparent increased functional connectivity in the EPT group in a contiguous band within the canonical beta frequency range (20-25 Hz) and in the canonical gamma frequency range (61-70 Hz). This hyperconnectivity was assessed and non-parametrically tested for group differences in network extent using NBS with 5000 permutations and familywise error correction set at p<0.05. A subnetwork was identified in the beta range (Figure 3) which was significant at a range of t statistic thresholds from 1 to 2. An addition subnetwork was identified in NBS within the low gamma range which was significant at a range of t statistic thresholds from 1 to 4 (Figure 4). Network strength was computed for both the beta and the gamma subnetworks and related to performance on standardized assessments of language and general abilities across and within groups.

**Figure 2:**
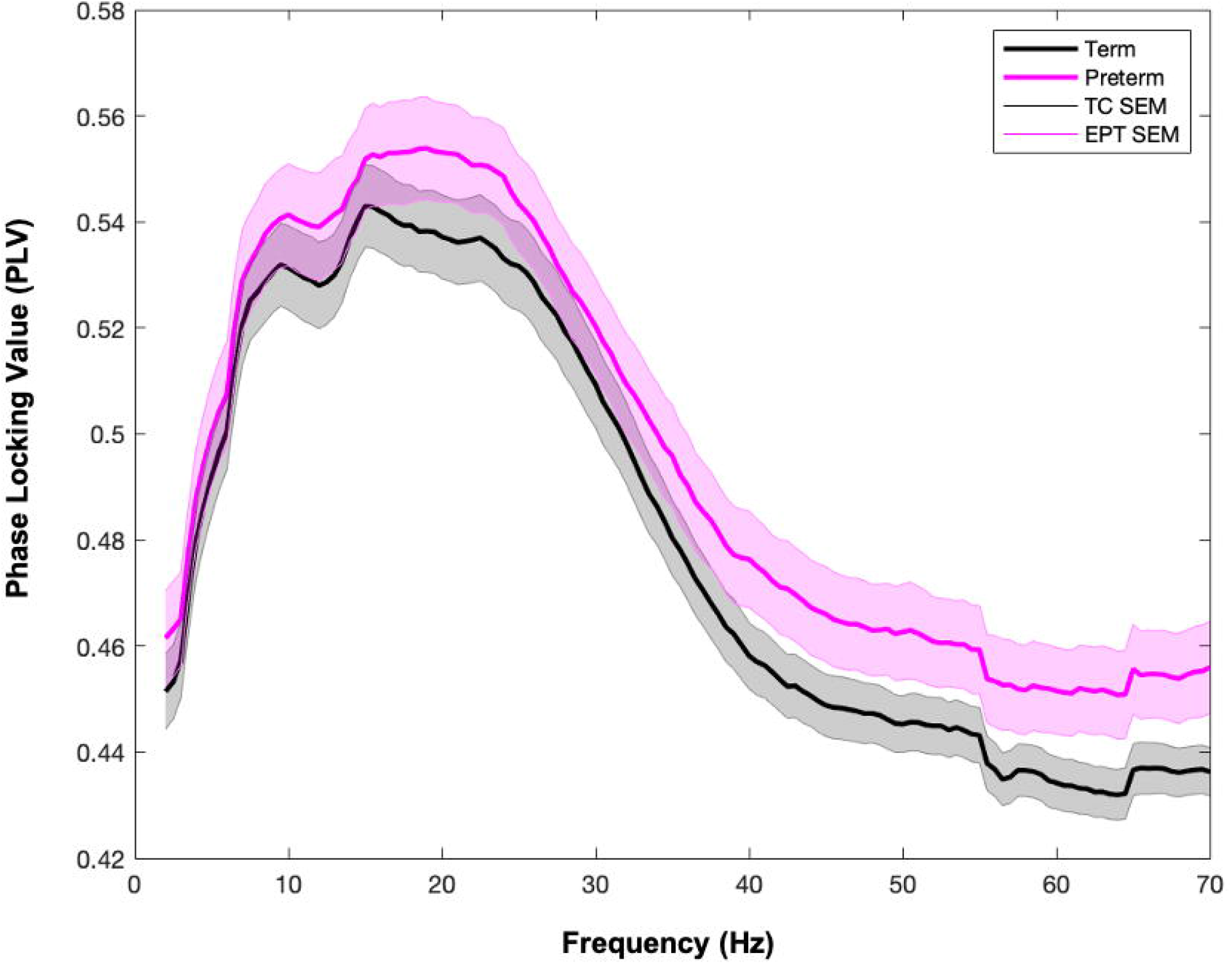
Functional connectivity indexed by phase locking value (PLV) for each group. Phase locking value (PLV) throughout the language network identified in Figure 1 for extremely preterm children (EPT, *n* = 31, shown in pink) and term controls (TC, *n* = 42, in black) with standard error shaded around the mean (SEM). Areas of group differences on visual inspection include segments of the beta band (20-25 Hz) and the gamma band (61-70 Hz).

**Figure 3:**
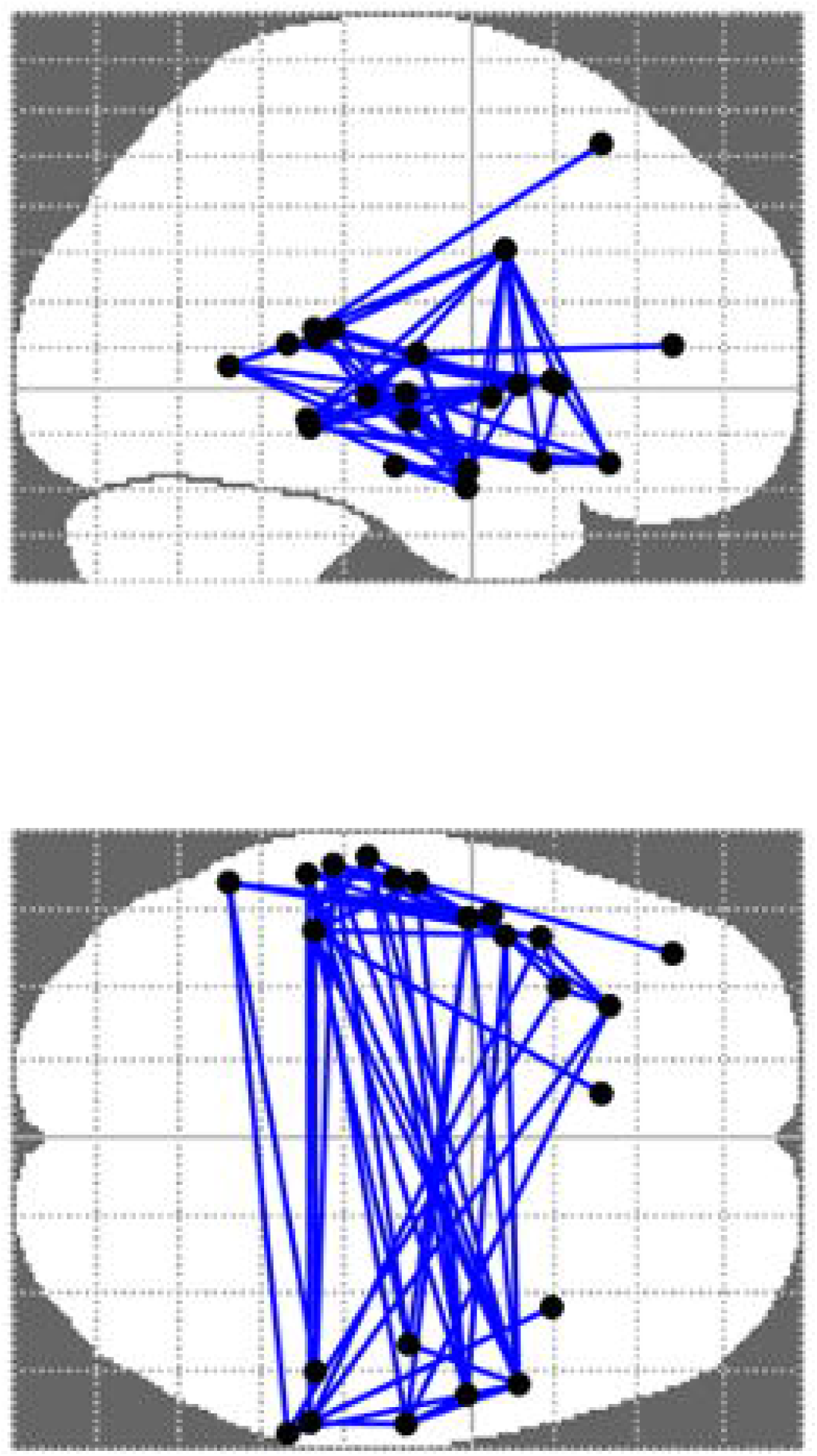
Extremely preterm children show increased beta functional connectivity in language network. Network “edges” showing significantly increased functional connectivity in EPT versus TC between 20 and 25 Hz during verb generation (observed at various initial thresholds ranging from *t* =1 to 2, 5000 iterations, *p* < 0.05, corrected for multiple comparisons).

**Figure 4:**
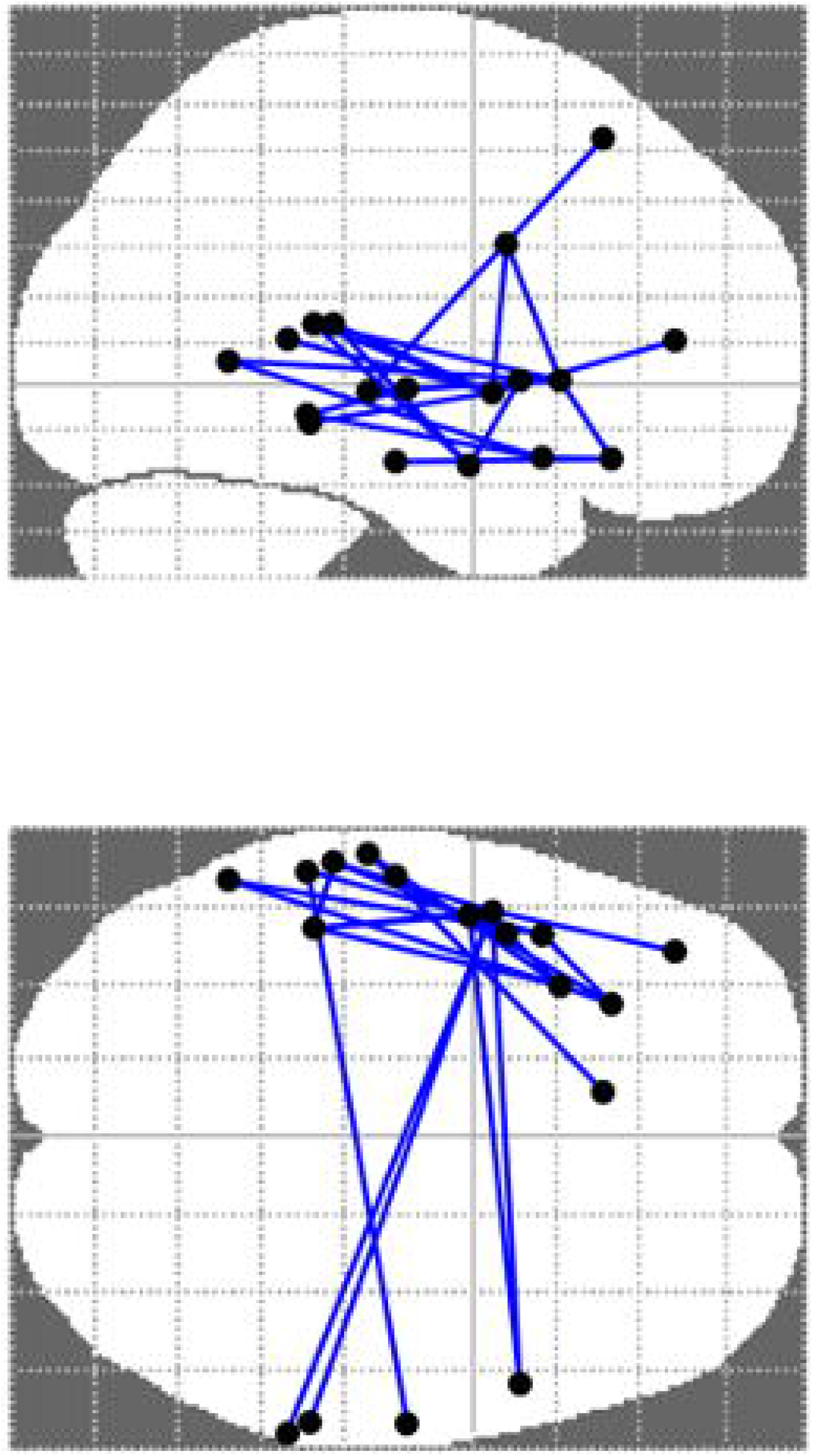
Extremely preterm children show increased gamma functional connectivity in language network. Network “edges” showing significantly increased functional connectivity in EPT versus TC between 61 and 70 Hz during verb generation (observed at various initial thresholds ranging from *t* =1 to 4, 5000 iterations, *p* < 0.05, corrected for multiple comparisons).

Strength in the beta and gamma subnetworks was not significantly related to performance across groups or within the TC group. However, for the EPT group exclusively, strength of the subnetwork identified in the gamma frequency band was positively correlated with standardized expressive language scores on the EVT (Figure 5, r = 0.32, p<0.05).

**Figure 5:**
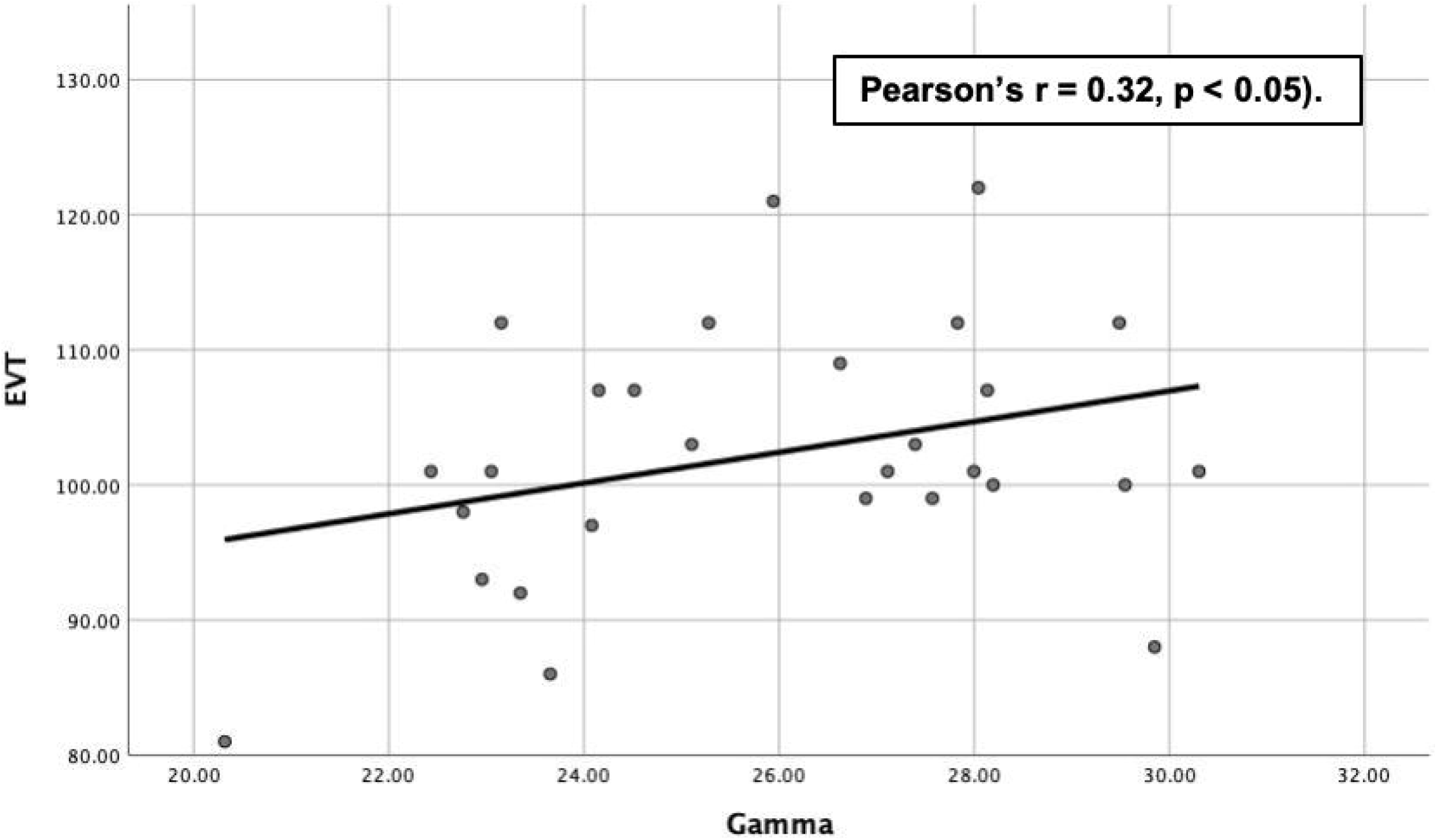
Relationship between gamma band-limited connectivity (network strength) in the identified subnetwork supporting language and expressive language scores for the preterm group. Scores on the Expressive Vocabulary Test (EVT2) versus network strength within the significant subnetwork identified in gamma band-limited connectivity for the EPT group exclusively. Results indicate a positive bivariate correlation (Pearson’s r = 0.32, p < 0.05). Correlations between EVT scores and network strength were not significant across groups or within the term control group.

As an exploratory analysis, we assessed for significant differences in language scores and brain connectivity by sex (female versus male) due to the known differential effect of sex on prematurity and language (Benavides et al., 2019; O’Driscoll, McGovern, Greene, & Molloy, 2018). Within the EPT group, there were no significant differences in neurobehavioral scores, but females did have significantly higher network strength in both the beta band (mean strength 65.39 for males and 70.89 for females, p < 0.05) and in the gamma band (mean strength 24.8 for males and 26.88 for females, p < 0.05). Across groups, there were no significant differences by sex in neurobehavioral scores or gamma connectivity, but there was significantly higher network strength in the beta band for females (mean strength 59.0 for males and 64.66 for females, p < 0.05). Within the control group, there were no significant differences between males and females.

## 4. DISCUSSION

In this contemporary cohort study of extremely preterm children with no known brain injury or neurological disorder and their term comparison children, we demonstrate significantly increased functional connectivity as indexed by MEG in preterm children versus term controls. This functional hyperconnectivity is significantly related to performance on standardized assessments of expressive language for the EPT group exclusively, despite no significant differences on language representation using conventionally analyzed task-based functional MRI. There were no significant differences between groups in WNV scores, used as a marker of general abilities. Functional connectivity was not significantly related to WNV scores, so our observed findings should not be attributed to global differences in performance or intelligence. As such, functional hyperconnectivity of areas known to support language in preterm children might represent an alternate developmental trajectory or compensatory network which enables EPT children to perform within normal limits on language assessments and comparably to term controls. This work is congruent with our earlier MEG and fMRI work on a smaller sample of EPT children and their term controls reporting functional and effective hyperconnectivity in the beta band for preterm children during a receptive stories listing task.

This work is significant in that it reports neuromagnetic findings in the largest cohort of extremely preterm children with magnetoencephalography and structural and functional magnetic resonance imaging that we know of to date. It is also important to note that these children are without known brain injury, so the findings likely represent a relatively “pure” effect of the dysmaturation that occurs due to preterm birth and not due to other common comorbid conditions such as periventricular leukomalacia or interventricular hemorrhage. There were no significant differences between groups in terms of family income or parental education. These variables are often used in indices of socioeconomic status, or SES, which are known to have differential impacts on both language and outcomes in prematurity. Furthermore, although there were no significant effects of sex on the connectivity results or on neuropsychological assessments when explored through bivariate correlations or independent samples t tests across groups (EPT + TC) or within the TC group, within the EPT group there were significant sex differences, with females exhibiting significantly increased network strength in both the beta and gamma frequency bands. There were no performance differences between preterm males and females. This finding warrants further investigation.

When viewed in the context of reported research from our group and from other labs, it seems that differential neuronal activation frequencies (beta versus gamma, etc.) and relationship to performance could be due to different task demands. For example, our prior work reported functional hyperconnectivity in the beta frequency range during a stories listening task. This verb generation tasks requires not only auditory processing and comprehension but also semantic retrieval and language production. The gamma hyperconnectivity we report as being significantly related to expressive language scores is known to be important in the typical development of language skills in children. Benasich et al found that children less than 3 years of age with a family history of language impairment had lower resting gamma connectivity on electroencephalography than term born children without a history of language impairment (Benasich et al., 2008). In prior EEG work investigating the ontogeny of EEG activity and synchronization before and after term equivalent age, gamma activation is of particular interest (Vanhatalo & Kaila, 2006). Higher frequency activation and synchrony becomes more prominent as interneuronal signaling matures during the same gestational period in which our preterm participants were born (Tokariev, Palmu, Lano, Metsäranta, & Vanhatalo, 2012; Vanhatalo & Kaila, 2006).

Our study has some relative limitations. The verb generation task was covert. Therefore, it might be difficult to gauge engagement and performance during the task. However, all participants demonstrated they could perform the task well during the practice items. Additionally, 11 out of 73 children had structural MRI and task-based MEG but did not complete the verb generation task in functional MRI. The verb generation task in fMRI was the last task of our study visit, and was sometimes excluded from the visit if the family was late for their appointment due to time constraints or stopped early if the child stated they were finished with imaging for the day. We will attempt to rectify this in future studies. We combined task-based fMRI data from 2 slightly different acquisitions on the same scanner using the same stimuli. We do not run connectivity analyses on the fMRI data and use it only to determine if differences in representation differ between groups before binarizing the first level contrast maps of activation to extract virtual sensors for MEG analyses. There were no differences in fMRI representation by group (both with and without regressing out activation differences due to acquisition). Therefore, differences in fMRI acquisition (if any exist) should not impact our MEG connectivity analyses. Finally, our preterm group and control groups both perform very well on standardized assessments of language (although the EPT group has significantly lower scores than the TC group). It is important to note that this is a community acquired self-selected sample and, therefore, might be higher performing than the general population. However, both groups perform within normal limits (standardized mean of 100 +/- 1 standard deviation) and our research question specifically pertains to factors that might help optimize performance in our clinical population.

Our study has a number of unique strengths. This is a contemporary cohort of extremely preterm children from a relatively narrow band of gestational ages (24-27 weeks), including some periviable children for whom resuscitation was entirely elective at time of birth. We excluded known comorbidities of prematurity, such as periventricular leukomalacia and neurological disorders. Due to differential rates of--and impact from-- prematurity, we assessed any differences in sex, race, ethnicity, and socioeconomic status and accounted for these factors in analysis, finding no significant differences. Finally, we used a multimodal neuroimaging pipeline that enables investigation into the cortical morphometry (Barnes-Davis, Williamson, Merhar, Holland, & Kadis, 2020a); structural connectivity (Barnes-Davis et al., 2020b); functional connectivity (Barnes-Davis et al., 2018); and neurobehavioral outcomes for our preterm and term participants at a single session. Multimodal investigations such as this have received attention recently in the scientific community as a way to increase reproducibility in functional imaging (Botvinik-Nezer et al., 2020).

In conclusion, we have demonstrated functional hyperconnectivity as indexed by fMRI-constrained MEG during a covert auditory verb generation task in the gamma frequency band for well-performing children born extremely preterm. Strength in this significant subnetwork (consisting of bilateral temporal and left frontal nodes) positively correlated with expressive language performance for the preterm group exclusively. This might serve as a marker of resiliency in preterm children with no known brain injury or neurological deficit. Future work will expand this cohort and follow them longitudinally to investigate the predictive power of our reported functional hyperconnectivity.

## Data Availability

De-identified data reported in this paper are available from the corresponding author (Dr. Barnes-Davis) on reasonable request.

## 5. ACKNOWLEDGEMENTS

We thank the participants and their families. We acknowledge the study coordination of Adrienne King, Cameron Laue, Claudio Toro-Serey, Sara Stacey, and Sandra Wuertz. This work was funded by an award from the National Institute of Child Health and Human Development (K12 HD028827 for MEB-D); an award from the National Center for Advancing Translational Sciences (KL2 TR001426 for SLM); awards from the National Institute of Neurological Disorders and Stroke (R01-NS096037 and R01-NS094200 for NAP); and by a Trustee Award Grant (DSK), a Shared Facilities Discovery Award (SLM), and an Arnold W. Strauss Fellowship Award for Excellence in Research (MEB-D) from the Cincinnati Children’s Hospital Medical Center.

## AUTHOR CONTRIBUTION STATEMENT

### Maria Barnes-Davis, MD/PhD

Conceptualization; Resources; Data curation; Formal analysis; Funding acquisition; Investigation; Methodology; Project administration; Roles/Writing - original draft; Writing - review & editing.

### Stephanie L. Merhar, MD/MS

Conceptualization; Resources; Funding acquisition; Investigation; Methodology; Supervision; Writing - review & editing.

### Scott K. Holland, PhD

Conceptualization; Supervision; Writing - review & editing.

### Nehal A. Parikh, DO/MS

Conceptualization; Resources; Funding acquisition; Investigation; Methodology; Supervision; Writing - review & editing.

### Darren S. Kadis, PhD

Conceptualization; Resources; Data curation; Formal analysis; Funding acquisition; Investigation; Methodology; Project administration; Supervision; Writing - review & editing.

